# Study Protocol for a Phase III Randomised Controlled Trial of Sailuotong (SLT) for Vascular Dementia and Alzheimer’s Disease with Cerebrovascular Disease

**DOI:** 10.1101/2022.03.24.22271670

**Authors:** Diana Karamacoska, Daniel K.Y. Chan, Isabella Leung, Jian-xun Liu, Henry Brodaty, Paul P Fahey, Alan Bensoussan, Dennis H. Chang

## Abstract

**Background:** Vascular dementia (VaD) accounts for 15-20% of all dementia cases. It is a syndrome of acquired cognitive impairment with a complex pathophysiological basis. A novel herbal formulation (Sailuotong; SLT) consisting of Panax ginseng, Ginkgo biloba and Crocus sativus extracts was developed to treat VaD. Preclinical animal studies found significant improvements in memory and in pathogenic biochemical parameters. Appropriate safety of SLT was shown in acute and chronic toxicity studies, and early clinical trials of SLT demonstrated enhancements in cognition in VaD patients. A fully powered study with a long intervention period is needed to confirm the efficacy and safety of this novel intervention.

**Methods:** A rigorous phase III clinical trial was developed with the aim of recruiting 238 patients diagnosed with mild to moderate probable VaD, or VaD mixed with Alzheimer’s disease (where cerebrovascular disease is the clinical dominant contributor to dementia, abbreviated as CVD+AD). Using a permuted block strategy, participants will be randomly allocated to receive SLT (120 mg bd) or placebo capsules for an intervention period of 52 weeks and will be followed-up for an additional 13 weeks. The primary outcome measures are the Vascular Dementia Assessment Scale-cognitive subscale and Alzheimer’s Disease Cooperative Study-Activities of Daily Living scale. Secondary outcome measures include the Clinician’s Interview Based Impression of Change-Plus, CLOX, EXIT-25, Neuropsychiatric Inventory-Clinician rating scale, and Dementia Quality of Life questionnaire. Safety is assessed through adverse event reports and liver, renal, and coagulation studies.

**Discussion:** Primary and secondary outcome measures will be compared between treatment and placebo groups, using intention to treat and per protocol analyses. We hypothesise that a 52-week treatment of SLT will be clinically effective and well tolerated in participants with VaD or AD+CVD. This project will provide vital efficacy and safety data for this novel treatment approach to VaD.

**Trial registration:** Australian New Zealand Clinical Trials Registry (ANZCTR), ACTRN 12616000057482. Registered on 20 January 2016. https://www.anzctr.org.au/Trial/Registration/TrialReview.aspx?id=369471&isReview=true

## BACKGROUND

Dementia is the leading cause of disability in older adults (1) and is the second leading cause of death in Australia.(2) Vascular dementia (VaD) is a clinical syndrome of acquired intellectual and functional impairment from conditions that damage blood vessels in the brain, such as stroke and cerebrovascular disease (CVD).(3) It is the second most common cause of dementia after Alzheimer’s disease (AD), accounting for ∼20% of all cases in western countries.(3) The prevalence of VaD is between 1 and 4% in individuals aged over 65 years,(4) and coexists with AD in ∼40% of cases.(5) Dementia significantly impacts on the life quality of patients and their caregivers, imposing a costly burden on the community and health care system. In 2017, the total cost of dementia (including VaD) to Australia was estimated at $14.67 billion, and this is anticipated to rise to $36 billion in the next 30 years.(6)

Currently, viable pharmaceutical options for VaD are lacking. Treatment largely focuses on symptom management and preventing further brain damage by controlling vascular risk factors such as hypertension and atherosclerosis.(7) Pharmaceutical agents developed to treat AD such as donepezil, galantamine, and memantine are also used by clinicians for managing symptoms associated with VaD (8). Although modest short-term clinical benefits for VaD were demonstrated in clinical trials, their usage is cautioned due to safety concerns, financial costs, and unknown long-term effects.(8–12)

Sailuotong (SLT) is a novel compound Chinese herbal medicine preparation that was developed to address the complex pathophysiology of vascular cognitive impairment.(13,14) The SLT formula consists of specific dosages of Panax ginseng C A Mey (ginseng), Ginkgo biloba L (ginkgo), and Crocus sativus L (saffron). The chemical profile of SLT was clearly defined using modern chemistry and pharmaceutical techniques, and 10 bioactive components were used as quality assurance markers for the final formulation of SLT (14). A series of *in vitro* and animal studies were conducted to evaluate SLT’s safety and mechanisms of action.(15–22) As summarised in Table 1, the bioactive components of SLT have demonstrated beneficial effects on multiple therapeutic targets associated with VaD.(14)

**Table 1.** The multi-targeting effects of SLT components on VaD pathophysiology.

| Therapeutic targets | Ginseng | Ginkgo | Saffron |
| --- | --- | --- | --- |
| Regulates amino acids | X | X |  |
| Anti-hypercalcemia | X | X | X |
| Anti-inflammatory | X | X |  |
| Anti-oxidant | X | X |  |
| Enhances cholinergic function | X | X |  |
| Anti-apoptotic | X | X |  |
| Enhances cytoskeleton function |  | X |  |
| Anti-thrombotic | X | X |  |
| Anti-platelet aggregation |  | X | X |
| Increases blood flow | X | X |  |

Three human clinical trials have demonstrated the therapeutic effects of SLT.(25–27) Neurocognitive enhancements were obtained in healthy adults,(25) and a pilot study with 62 VaD patients found SLT improved ADAS-cog scores and quality of life, with a subset of 17 patients showing increased cerebral blood flow to the frontal and temporal lobes of the brain.(16,26) A recent phase II clinical trial, with 325 VaD patients recruited in China, reported significant enhancements in memory, executive function, language, and orientation following 26 weeks of SLT treatment.(27) Safety and tolerability of SLT were also demonstrated in these trials. One in 10 participants experienced stomach discomfort, rash, diarrhoea, heartburn, insomnia, headache, and abnormal alanine aminotransferase, aminotransferase and thrombin time in laboratory results; all were considered mild reactions that ceased when the SLT was stopped.(27,28) Serious adverse events (SAEs) were reported in one trial, however, these were all deemed to be unrelated to SLT.(27) Acute toxicity studies also found no significant herb-drug interactions.(19) Together, this research demonstrates that this novel intervention may be effective and safe for VaD. The present phase III clinical trial aims to further evaluate the efficacy and safety of SLT using a longer intervention period (52 weeks) in an Australian cohort.

## METHOD

### Study aims

The primary aim of this research is to assess the effectiveness of a 52-week treatment with SLT on cognition (Vascular Dementia Assessment Scale-cognitive Subscale; VaDAS-cog) and functioning (Alzheimer’s disease Cooperative Study-activities of daily living scale; ADCS-ADL) in people diagnosed with mild to moderate VaD or AD+CVD. The secondary aims of the study are to evaluate executive functioning (EXIT-25 and CLOX), mental health (NeuroPsychiatric Inventory; NPI), and the clinician’s impression of change (CIBIC-plus), and review the participant’s quality of life (Dementia Quality of Life questionnaire) from the perspective of the participant and their caregiver. Safety will be determined with adverse event (AE) reports and laboratory studies.

### Study design and setting

This study is a multicentre phase III two-arm randomised, double-blind, placebo controlled clinical trial of 65 weeks, including a 52-week intervention and a 13-week follow-up. Eligible participants are randomized into one of two parallel treatment groups: the Intervention group taking SLT (Active) or Placebo control group (Placebo). Participants and investigators (including persons responsible for data collection, data management and data analysis) will not be aware of randomisation assignments. The study is being conducted across several hospitals and clinics in Australia.

### Participant characteristics

A broad representation of Australian adults according to demographic and social factors will be sought to improve the generalisability of the study findings for the community. As VaD and AD often coexist,(5) up to a third of participants with a mixed pathology (i.e., AD+CVD) will be included in the study. Participants with a confirmed diagnosis of probable VaD will still constitute at least two thirds of the total sample.

#### Inclusion criteria

Volunteers will be assessed for study eligibility using the following inclusion criteria:

- Aged 40–85 years old*;
- Confirmed diagnosis of probable VaD, as defined by the National Institute of Neurological Disorders and Stroke (NINDS) and the Association Internationale pour la Recherche et l’Enseignement en Neurosciences (AIREN),(29) or possible Alzheimer’s Disease with significant neuroimaging (CT or MRI) evidence of cerebrovascular disease, as defined by the National Institute of Neurological and Communication Disorders and Stroke (NINCDS)-Alzheimer’s Disease and Related Disorders Association (ADRDA)(30);
- Score between 10–24 on the Mini Mental State Exam (MMSE) for the diagnosis of mild to moderate dementia;
- Score ≤ 11 on the Geriatric Depression Scale (GDS) for the absence of severe depression;
- Stable or controlled by optimal medication over a minimum of 6 months if on ChE inhibitors and 3 months for (if present) hypertension, diabetes, cardiac disease or stroke, or if on hypnotics and sedatives, stabilised for more than 3 months prior to inclusion in the study;
- Agreement to take part in the study as evidenced by a personally signed and dated informed consent document indicating that the volunteer (or a legally acceptable representative if the participant is unable to provide consent) has been informed of all pertinent aspects of the study;
- Participants must also be accompanied by a caregiver, and this person must be able to assist the participant comply with the study protocol. This requires the caregiver to be in contact with the participant at least 2 days per week;
- If female, has no intention to become pregnant during the study.

* Rule waiver: a participant aged over 85 years may still be eligible for enrolment if he/she fulfils the following two conditions:

1. the participant meets all inclusion and exclusion criteria of the study;
2. participant’s health status is likely to allow for successful completion of the 15-month study based on clinical judgement of the Principal Investigator (PI) after assessing the participant’s medical history, current health and comorbidities and factors potentially affecting compliance (e.g., availability of a reliable caregiver). A 9-point Clinical Frailty Scale will be used to facilitate the PI’s decision-making process. This scale is a well validated tool used clinically to predict death or need for institutional care in the elderly.(31) The participant may be considered suitable for this study if his/her frailty score is less or equal to 5 (mildly frail). In such a scenario, a rule waiver will be completed before enrolment.

#### Exclusion criteria

Participants will be excluded from the trial if they:

- Have other types of dementia and/or severe form of delirium, depression, schizophrenia, acute illness or poorly controlled chronic diseases;
- Receive administration of the ingredients in the SLT formula (ginseng, ginkgo and saffron);
- Have a history of severe forms of peptic ulcers, diabetes with complications, pulmonary disorders, renal and/or hepatic disorders;
- Have had a stroke in the 3 months before screening;
- Have abnormal pathology test results: Creatinine > 1.5 times upper limit of normal (ULN); Aspartate transaminase, Alanine transaminase or Alkaline Phosphatase > 2 times ULN; Prothrombin time > 3 seconds more than ULN; Activated Partial Thromboplastin Time > 10 seconds more than ULN; Platelet count < 100×10^9^/L;
- Have severe dysphasia or mental retardation;
- Have life expectancy < 6 months;
- Are allergic to more than 2 medications or at least 1 ingredient of SLT;
- Are participating in another clinical trial;
- Are pregnant or lactating.

### Intervention

The active (SLT) and placebo preparations will be manufactured in a facility certified with Good Manufacturing Practice by the Australian Therapeutic Goods Administration.

#### Active SLT treatment

Each SLT capsule contains 60 mg of standardised bioactive extracts including 27.27 mg of Ginsenosides from *Panax ginseng*, 27.27 mg of Ginkgo flavone glycosides from *Ginkgo biloba*, and 5.46 mg of Crocins from *Crocus sativa*. The treatment dose of SLT is 240 mg daily (4 × 60 mg capsules – 2 in the morning and 2 in the evening) of SLT daily for 52 weeks.

#### Placebo

Placebo capsules will be created with an inert substance (starch filler, silica coating) and matched with SLT for colour, taste, texture, and weight. Participants in this group will also take 2 capsules, each morning and evening, for 52 weeks.

#### Discontinuation of participants from treatment or assessment

Should any of the exclusion criteria present after enrolment of the study, the Investigator will be required to immediately contact the Coordinating Chief Investigator to determine the next appropriate course of action. Participants who finish their baseline examination and begin taking the blinded medication will be counted as withdrawal cases if, during the study they:

- Become pregnant;
- Withdraw informed consent. Participants are free to discontinue their participation in the trial at any time, without prejudice to further treatment;
- Become lost to follow-up during the 52-week trial period;
- Demonstrate significant protocol non-compliance as determined by the Investigator; or
- The Investigator considers that it is not in their interest to continue the study.

The study may be discontinued at any time by the Sponsor or the Coordinating Chief Investigator based on new information regarding safety or efficacy. Additionally, the study may be terminated if progress is unsatisfactory. Withdrawn/discontinued participants will not be replaced.

#### Procedures for discontinuation

In case of premature termination or suspension of the trial, the Coordinating Chief Investigator must inform the trial participants and ensure appropriate follow up and therapy. In addition, the regulatory authorities and ethics committee must be informed.

Participants may withdraw/discontinue from the study at will, at any time without explanation. The participant may also be discontinued at the discretion of the Investigator due to a safety concern or if judged to be non-compliant with trial procedures. Participants that withdraw/discontinue from the study will be asked to attend the end of treatment visit (Week 52) to monitor their safety, within 14 days of their withdrawal. The data collected up until the withdrawal visit will be retained to ensure that the results of the research project can be measured properly. Participants must advise the researchers if they wish to have their data removed from the project at the time of their withdrawal.

#### Compliance and success of blinding

On an ongoing basis, all study drugs will be reconciled against delivery, use, and returned medication documents. The number of returned capsules will be counted by the delegated research personnel at each site. To be compliant with the treatment protocol, study drug dosing must have a compliance rate >70% over the 52-week trial period. Self-reported measures of compliance will involve asking participants and their caregiver to complete a diary between visits. The researchers will also ask about their adherence to the prescribed treatment regime, and which treatment condition they think that they were allocated (with response options of SLT, placebo, or unsure), at the week 4, 13, 26, 39 and 52 visits.

#### Concomitant treatment

Participants may take routinely prescribed medications provided the relevant condition has been stable or controlled by optimal medication for more than 3 months except those drugs prohibited by the Exclusion Criteria. For anti-AD medications (e.g., ChE inhibitors), the doses should be stable for at least 6 months prior to the commencement of the trial and remain stable for the duration of the study. The details of all medical treatments will be recorded as part of the screening process and then, following enrolment, during every follow-up visit. It is anticipated that any unmeasured confounders or effect modifiers will be equally distributed between groups due to the randomisation process.

### Primary Outcome Measures

There are two co-primary outcome measures in this study:

- The Vascular Dementia Assessment Scale-cognitive subscale (VaDAS-cog) will be used as the primary outcome measure of cognitive function in this trial. VaDAS-cog is the modified version of the Alzheimer’s Disease Assessment Scale cognitive subscale (ADAS-cog).(32) It is a sensitive psychometric scale for assessing the severity of cognitive impairment over time in dementia participants and covers four core symptoms of dementia: memory, orientation, language, and praxis. VaDAS-cog comprises additional frontal lobe subtests covering attention, working memory, executive function and verbal fluency to reflect the unique pathological feature of VaD.(33)
- The Alzheimer’s Disease Co-operative Study-Activities of Daily Living inventory (ADCS-ADL) is a validated instrument for determining the level of functional disability in dementia patients.(34) The scale is comprised of basic activities (eating, walking, toileting, bathing, etc) and complex, instrumental activities (using the telephone, preparing a beverage or meal, using household appliances, etc).

### Secondary Outcome Measures

- Global assessment of change will be measured by the Clinician’s Interview Based Impression of Change-plus (CIBIC-plus). This is a reliable tool to rate the participant’s condition by a clinician experienced in managing patients with dementia (global assessment).(35) CIBIC-plus is a semi-structured interview that includes four categories for evaluation – general, mental/cognitive state, behaviour and activities of daily living.
- Small vessel disease that causes subcortical ischemic impairments, including lacunar infarcts and ischaemic white-matter lesions, is the most common type of VaD in elderly people,(36) contributing to executive dysfunction. Additional executive function tests will be performed using CLOX and EXIT-25. Both instruments have been validated in various dementia cohorts including VaD.
  – The CLOX is a modified version of a clock drawing task that is a quick and sensitive tool for the assessment of executive function in elderly dementia patients. It has also been used as a rapid method for dementia screening, as it shows strong correlations with other traditional cognitive measures.(37)
  – The EXIT-25 was developed by the same group of researchers who developed CLOX, with the aim of defining the behavioural sequelae of executive dysfunction in dementia patients. EXIT-25 correlates well with other executive control function tests, such as the Wisconsin Card Sorting Test.(38,39)
- Quality of life is assessed through the DEMQOL, a dementia-specific Health-related Quality of Life (HRQOL) instrument. DEMQOL has both self-(28-item) and proxy-rating (31-item) components providing different but complementary perspectives, from the participant and their caregiver, on quality of life with dementia. DEMQOL also shows comparable psychometric properties to the best available dementia-specific measures.(40)
- Neuropsychiatric Inventory-Clinician rating scale (NPI-C) is a revised version of the Neuropsychiatric Inventory (NPI). The latter is a popular tool for evaluating neuropsychiatric symptoms of delusions, hallucinations, depression/dysphoria, anxiety, agitation/aggression, euphoria, disinhibition, irritability/lability, apathy, aberrant motor activity and night-time behaviour disturbances, based on a structured interview with a caregiver.(41) The NPI-C has a clinician-rating methodology, that consistently demonstrates high inter-rater reliability and convergent validity in dementia participants.(42)

### Safety Measures

Safety is evaluated through laboratory studies and AE reports. Routine tests of haematology (full blood count), coagulation (Prothrombin time, Activated Partial Thromboplastin Time, and Fibrinogen Level), liver function (Total Bilirubin, Aspartate transaminase, Alanine transaminase, Alkaline Phosphatase, Albumin, and Total Protein), and renal function (Blood urea nitrogen, Creatinine, and Sodium) will be conducted at the Screening visit and monitored for changes 4, 13, 26, 39 and 52 weeks after treatment is commenced. AEs, and any worsening of symptoms, will be closely monitored throughout the study. The relationship between abnormal blood test results or AEs and the treatment will be determined by the Investigators and/or trial physicians. If a serious adverse event occurs that is unexpected, and the treatment is suspected as the cause, the randomisation code may be broken to assist in the decision-making process for providing appropriate medical care. An external university staff member, who generated the randomisation sequence, will carry out the unblinding process and subsequent reporting to the Sponsor and regulatory bodies.

### Study Protocol

Figure 1 details the assessment schedule. The following section describes the procedures for recruitment, screening, randomisation, treatment and assessment, and review of safety and laboratory studies. A schematic of the study protocol is shown in Figure 2.

**Figure 1.**
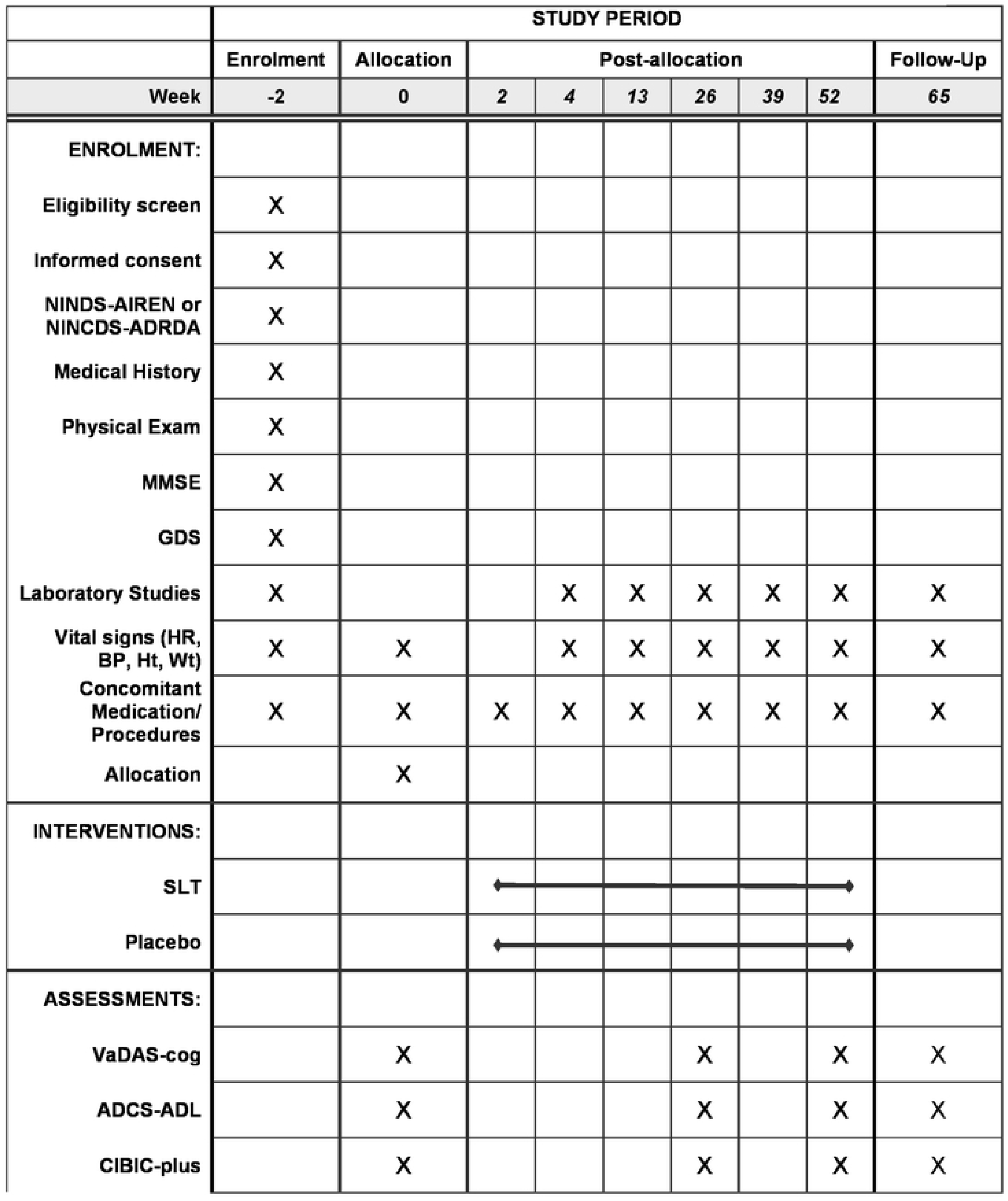
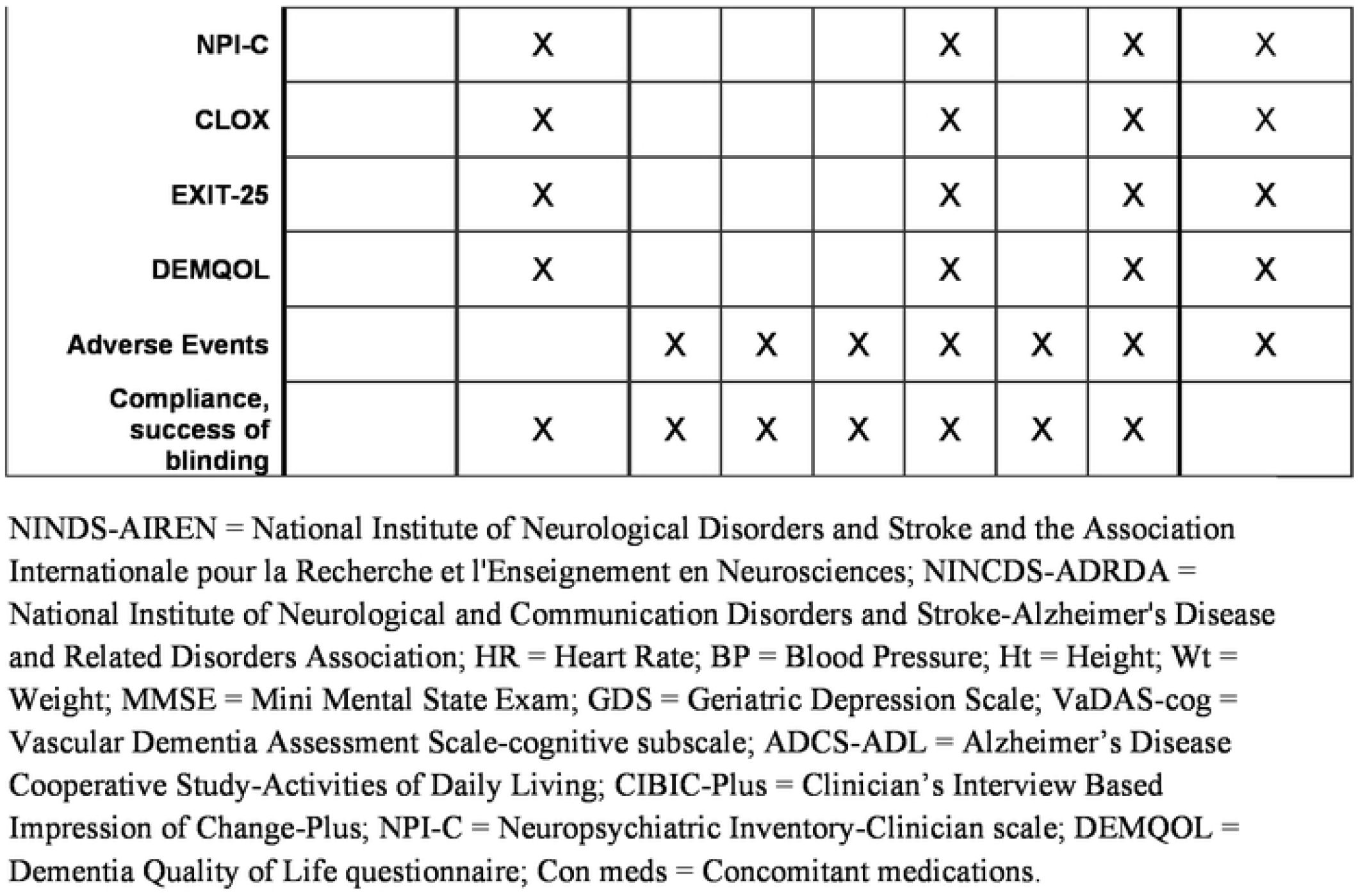
Schedule of enrolment, assessments, and timepoints.

**Figure 2.**
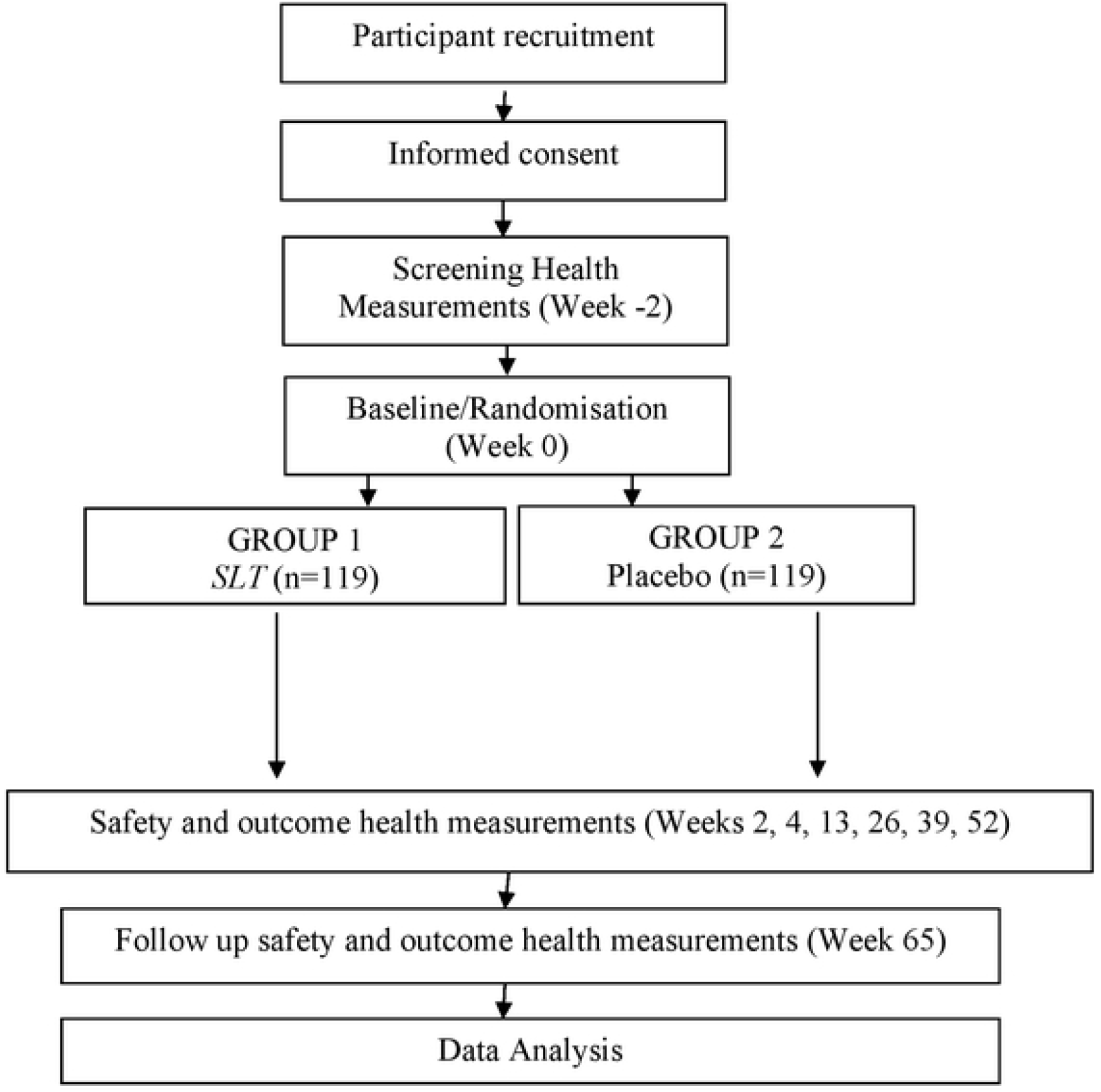
Schematic of the Trial Protocol.

#### Screening

At the screening visit, volunteers and their caregiver will have the nature, purpose, and risks of the study explained to them by the Investigator. Written informed consent will be obtained from the participant (or their legally acceptable representative), and their caregiver. The participant will be assessed for eligibility by the Investigator. Medical history, prior and concurrent medications, planned procedures, and demographic information will be documented, in addition to completing a physical examination, having body weight, height, and vital signs measured, and having blood collected and analysed. The screening visit should occur within 2 weeks prior to randomisation (week 0).

Volunteers who are found to be ineligible for the study will be notified of this outcome and any abnormal findings/test results. Their regular doctors may also be contacted if the nature of these findings warrants this.

#### Treatment protocol and assessment schedule

Eligible participants will be asked to return for a baseline exam of primary and secondary outcome measures. All data collected between weeks -2 (screening) and 0 (baseline) will be considered as baseline assessments for the study. The first 13 weeks of medication will be dispensed with instructions provided to take two capsules, orally, twice per day. After the commencement of treatment, progress and/or AE reporting will be conducted at weeks 2 (via phone), 4, 13, 26, 39, 52 and 65 (follow-up). The next supply of the 13-week medication is dispensed at week 13, 26 and 39 visits. An additional week of medication (14 weeks total) will be provided with each dispensing in case there are any unforeseen delays with the scheduled appointments. Primary and secondary outcome assessments are repeated at weeks 26 (midpoint) and 52 (endpoint). Follow-up assessments of the primary outcome measures and the clinician’s impression of change will be completed after the treatment period (week 65 visit) and any ongoing AEs will be closely monitored. Participants are required to keep a diary between all visits to monitor compliance, changes in medications, and AEs. Participants will be offered reimbursement to cover travel expenses at each visit.

#### Laboratory studies and AEs

Pathology samples will be collected and tested by local certified pathology laboratories. These studies will be conducted at the screening, and week 4, 13, 26, 39, 52 and 65 visits. Participants will need to refrain from caffeine and smoking on the morning before sample collection, and avoid alcohol and exercise for 24 hours prior to testing. Standard blood tests will be conducted for haematology, coagulation, liver function, renal function, and electrolytes (Potassium and Chloride). All clinically important abnormal laboratory results found during the study will be investigated until resolved.

Information about AEs will be collected at each visit, and participants can contact the PI about any concerning health problems, at any time. The Investigators will contact the research physicians and/or other members of the research team, who will review the dosage and continuation status in the trial, provide medical advice, and/or communicate with their General Practitioner or other health professionals, and coordinate emergency medical attention as is required. An AE will be followed up until it resolves or for up to a month after the study is concluded. The reporting of AEs will be consistent with TGA guidelines, which are in turn based on the Note for Guidance on Good Clinical Practice (CPMP/ICH/135/95) and Note for Guidance on Clinical Safety Data Management: Definitions and Standards for Expedited Reporting (CPMP/ICH/377/95),(43) and according to any requirements of the Human Research Ethics Committee.

#### Recruitment

Recruitment strategies to be used in the Australian sites will include:

- All certified geriatricians and neurologists involved in this trial and their associated hospitals, clinics and networks
- Advertising through Dementia Australia’s website
- Visits by the Investigators to major aged care outpatient and nursing home facilities
- Visits to Division of General Practice meetings/events
- Posters and brochures in aged care, general practices and other health facilities
- Recruiting through Step Up For Dementia Research website
- Media resources of Western Sydney University e.g. advertisements in local newspapers
- Advertising through newsletters and websites of NICM Health Research Institute, Western Sydney University
- Advertising through online social media platforms (e.g., twitter, Facebook, Instagram).

#### Randomisation and allocation concealment

The randomisation schedule was created by a university staff member, external to the investigative team, using a computer-generated randomisation sequence in Microsoft Excel. Randomisation numbers are allocated in permuted blocks of six, with an allocation ratio of 1:1. Allocation is concealed using batch numbers computer-generated by the same staff member; these were sent directly to the manufacturer. Medication containers will be labelled by the manufacturer, according to the randomisation schedule. Sites will allocate the randomisation numbers in order of number sequence starting with the lowest number in each block and using all numbers in a given block. Participants, caregivers, and all research personnel will be blinded to treatment allocation until the end of the trial, when data analyses are completed and the coding is unlocked, except in the case of emergency.

### Data Management

In accordance with ethical guidelines, the anonymity, confidentiality and privacy of participants will be protected. Source data records will capture screening and recruitment logs, drug accountability logs, informed consent documentation, clinic notes, expenditure records, participant case record forms (these records will be dated and signed by the PI) and AE reports forms. Hardcopy source data records will be kept in a locked cabinet in the office of the PI at each recruitment site during data collection. De-identified case record form data is transcribed to an electronic clinical trial management data system that is password protected. Western Sydney University will keep source or certified copies and electronic data for an indefinite period of time. All publication material will refer to general trial results as aggregate data and no individual participant names or identifying information will be released.

#### Monitoring

All research personnel will be trained in trial standard operating procedures and the use of the assessment tools. On-site and remote monitoring visits are conducted with each centre to review consent forms, case report forms, drug accountability records and investigator site files to verify compliance with Good Clinical Practice and the study protocol.

#### Data and Safety Monitoring Board

The Data and Safety Monitoring Board (DSMB) is responsible for monitoring safety data, trial progress and protocol adherence. The DSMB meets twice a year, or more often if required. After each DSMB meeting, a recommendation to continue, modify, or prematurely terminate the trial is made.

### Statistical Analysis

#### Sample Size

Following the release of the Phase II SLT trial results,(25) sample size for this study was recalculated as the two studies are based on similar participant cohorts and study design. In particular, the Phase II study provided useful data for VADAS-cog, one of the primary outcomes for the current study. Sample size was calculated using the Phase II trial outcome data for the two primary measures (VADAS-Cog and ADCS-ADLs). Based on their VADAS-Cog data,(25) we expect the minimum between-group endpoint difference to be 3 or more units (with a standard deviation of 6.56) for a clinically meaningful change in cognition. Using a one-sided independent samples t-test for the purpose of calculating sample size, a total of 166 participants are required to detect this difference with 90% power at the 0.05 significance level. For the ADCS-ADLs, we anticipate the minimum between-group endpoint difference to be 3 or more units (with a standard deviation of 5.31) for a clinically meaningful change in daily functioning. Using a one-sided independent samples t-test for the purpose of calculating sample size with this measure, a total of 110 participants are required to detect this difference with 90% power at the 0.05 significance level. Allowing for a total non-compliance/withdrawal rate of 30%, 238 participants will be recruited into the trial across all centres to ensure 166 participants in the final analyses.

#### Preliminary analyses

At completion of data collection, all variables and all logical pairs of variables will be subject to descriptive analyses using graphs, frequency counts and summary statistics. This will allow a) identification of unusual or unexpected results for data checking; and b) familiarisation with the distributions and associations within the data set. Where appropriate, variables which have non-symmetric distributions will be either transformed or categorised.

To address withdrawals, loss to follow-up or non-compliance with the study, analyses will be conducted on both an intention to treat (ITT) and per protocol (PP) basis. Withdrawing and non-compliant participants shall be encouraged to continue with data collection even if stopping treatment. Where data items are missing, the last value carried forward method to replace missing data in the ITT analysis will be used. Participants who have significant deviations from the protocol will be removed from the PP analysis after the completion of the ITT analysis. Such significant deviations from the protocol will be determined and documented by the study clinician during the study. Any deviation from randomisation, missing data and withdrawals will be fully reported for this purpose. In the case of death, all measurements prior to death will be included in the analysis but all after death will be set to missing. At the completion of data checking and correction, the data set will be locked for analysis using SAS and/or SPSS software.

#### Checking for homogeneity of study centres

As this is a multi-centre study, early analyses will address the question of whether there is heterogeneity between centres. Linear models will be fitted to each outcome measure in turn, with centre and centre by treatment added as fixed effects. Any statistically significant differences will be documented and explored further for potential confounding demographic or medical history factors. If variation between centres cannot be explained (or is shown to be related to differences in study methods), the primary analysis will continue as planned, but will be followed by sensitivity analyses which will either stratify by or exclude the outlier site(s) (depending on the sample size of the site[s] involved).

#### Demographic and baseline characteristics

The demographic and medical characteristics of participants in each treatment group will be summarised using percentages or means and standard deviations. Pearson’s Chi-square and independent samples t-tests will be used to check for any statistically significant differences between groups. Results will be documented as p-values and, where necessary, addressed within the interpretation of study results.

#### Primary efficacy analysis

The primary analysis will be linear mixed models through which we will test for differences between treatment groups on each outcome over the 52-week intervention period, with adjustment for random variation between treatment centres, with and without adjustment for other potentially important predictors (e.g. compliance, age, gender, severity of VaD). Non-linear changes over time will be tested with logistical regression by a) fitting time as a categorical variable; and b) testing for quadratic and cubic effects. Results will be reported as regression coefficients (or odds ratios for categorical variables) and associated 95% confidence intervals. Secondary analyses looking across time points will have greater power as more data points are included.

#### Subgroup analyses

Secondary analyses will be a repeat of the above stratified by disease type and, if necessary, with stratification by research centre.

#### Safety analyses

There will be no interim analyses apart from monitoring safety variables. The frequency, type and probable association of AEs will be tallied by de-identified treatment group every 3 months for review by the study team. If there is any significant safety issue identified, a meeting with the DSMB will be called.

## DISCUSSION

Based on prior clinical studies, SLT appears to be effective in enhancing neurocognition in healthy adults, and in improving cognitive functioning and cerebral perfusion in individuals diagnosed with VaD. These investigations have also shown SLT is well tolerated and considered safe in these cohorts, but the long-term outcomes have yet to be identified. The present phase III clinical trial will therefore establish vital efficacy and safety data for this novel standardised formulation. To address the issue relating to the common coexistence of VaD and AD(5), we include up to 30% of participants who have mixed vascular and AD pathology in this trial. In doing so, the generalisability of the study results are enhanced.

A rule waiver for participants aged over 85 years was implemented in this study, which impedes recruitment and the generalisability of our findings. This was a risk mitigation strategy we adopted due to the high incidence of death in older Australian cohorts. Around 70% of all dementia-related deaths occur in individuals aged 85 years or over.(2) To ensure their safety and suitability for this study, they are subject to additional evaluations by the Investigator. Recruitment difficulties have also arisen given the lower incidence rate of VaD amongst the population, relative to AD. This led to the expansion of this study to other Australian capital cities and the recruitment period being extended. However, the multi-centre aspect of this study poses several operational challenges. The identification of suitable sites requires a thorough evaluation of the site’s capacity to undertake a project of this size and the feasibility in recruiting the target population from the local area; a process that can take considerable time. Ensuring inter-rater reliability between- and within-study centres is also imperative. Thorough training on the trial instruments is provided at the site initiation visit and this is annually refreshed to ensure quality data collection. With the addition of trial sites and research personnel, monitoring efforts were increased for quality assurance purposes. This allows for the immediate identification and handling of any deviations or issues and minimises the heterogeneity between centres.

Due to the long duration of the study, and time spent at baseline, midpoint and endpoint visits, participant and caregiver burden is another concern. To overcome this, expectations will be explained prior to and at the screening visit, with reminders provided between visits regarding the type and duration of upcoming appointments. Breaks are also offered to reduce fatigue at these visits. Initiatives to enhance recruitment and retention for this study include reimbursing participants at each visit, and the provision of a 12-month supply of SLT at the conclusion of the 65-week study. This supply is subject to approvals from the Investigator and Coordinating Chief Investigator, with the participant’s (or a legally acceptable representative’s) agreement to being remotely monitored for their safety during this time.

## Data Availability

No datasets were generated or analysed during the current study. All relevant data from this study will be made available upon study completion.

## ABBREVIATIONS

VaD: Vascular dementia
AD: Alzheimer’s Disease
CVD: Cerebrovascular Disease
CVD+AD: Cerebrovascular Disease mixed with Alzheimer’s Disease
SLT: Sailuotong
NINDS-AIREN: National Institute of Neurological Disorders and Stroke and the Association Internationale pour la Recherche et l’Enseignement en Neurosciences
NINCDS-ADRDA: National Institute of Neurological and Communication Disorders and Stroke-Alzheimer’s Disease and Related Disorders Association
HR: Heart Rate
BP: Blood Pressure
Ht: Height
Wt: Weight
MMSE: Mini Mental State Exam
GDS: Geriatric Depression Scale
VaDAS-cog: Vascular Dementia Assessment Scale-cognitive subscale
ADCS-ADL: Alzheimer’s Disease Cooperative Study-Activities of Daily Living
CIBIC-Plus: Clinician’s Interview Based Impression of Change-Plus
NPI-C: Neuropsychiatric Inventory-Clinician scale
DEMQOL: Dementia Quality of Life questionnaire
Con meds: Concomitant medications
AE: Adverse Event
SAE: Serious Adverse Event
ULN: Upper Limit of Normal
TGA: Therapeutic Goods Administration
PI: Principal Investigator
DSMB: Data and Safety Monitoring Board

## DECLARATIONS

### Trial Status

Recruitment for this trial began on 01 August 2016 and recruitment is currently ongoing.

### Ethics approval and consent to participate

Ethical approval for this protocol (version 11) was requested and approved through the South West Sydney Local Health District (Approvals 2019/ETH08735 and HREC/14/LPOOL/81) and Western Sydney University (Approval H11554) Human Research Ethics Committees. All participants will provide informed consent before any study-related activities are undertaken. This study protocol was designed in line with Standard Protocol Items: Recommendations for Interventional Trials (SPIRIT) (see Additional file 1) and Good Clinical Practice guidelines. Any changes to the study protocol will be communicated with the approving ethics committees, study investigative team, and Australian New Zealand Clinical Trials Registry.

### Competing interests

As a medical research institute, NICM receives research grants and donations from foundations, universities, government agencies, individuals, and industry. Sponsors and donors provide untied funding for work to advance the vision and mission of the institute. The project that is the subject of this article was not undertaken as part of a contractual relationship with any organisation other than the funding that was previously declared in the Funding section. The authors declare that they have no competing interests.

### Funding

Australia Shineway Technology Pty Ltd provide the funding and medication to support the conduct of this trial. The Sponsor will in no way have access to or influence the data from this project.

### Authors’ contributions

DHC and AB conceptualised the study and its design, with input from DKYC, JL, PPF, and HB. DK drafted the manuscript for this study protocol, with assistance from IL, and all authors provided critical feedback. All authors read and approved the final manuscript.

## Acknowledgements

The authors would like to thank Mayryl Duxbury, Kellie Bilinski, Lena Hattom, and Genevieve Steiner for assisting with this project.

## Notes

### Competing Interest Statement

The authors have declared no competing interest.

### Clinical Trial

ACTRN12616000057482

### Funding Statement

Australia Shineway Technology Pty Ltd provide the funding and medication to support the conduct of this trial. The funders had and will not have a role in study design, data collection and analysis, decision to publish, or preparation of the manuscript.

### Author Declarations

Ethics committee/IRB of South West Sydney Local Health District and Western Sydney University gave ethical approval for this work

